# Patient-centred support for retinal diseases: Introduction of a telephone-based patient support programme

**DOI:** 10.1101/2025.11.05.25339217

**Authors:** Kai Januschowski, Eva-Maria Konrad, Anna Thonet, Florian Karsch, Virginia Ziska, Kristina Gelblin

## Abstract

**Background:** Patients with retinal diseases and their relatives face many challenges that may negatively impact treatment adherence that is required to maintain visual acuity. Patient support programmes (PSPs) may contribute to improve the understanding of the disease/therapy and promote intrinsically motivated treatment adherence.

**Methods:** In September 2023, a PSP was established for patients with neovascular age-related macular degeneration (nAMD), diabetic macular oedema (DME) or macular oedema due to retinal vein occlusion (RVO) and their relatives. As part of the PSP, each participant is individually supported by a personal contact through regular support calls. To gain an increasingly better understanding of patient needs, reactively collected health- and therapy-related data is stored in an anonymised manner.

**Results:** Since the start of the PSP, a total of 207 participants have registered for the PSP, with the majority being patients with nAMD (77.8%). The main topic of interest among patients and relatives was the diagnosis of nAMD (25.4% and 26.4% of respective inbound calls). Key patient topics also included treatment-related aspects such as application intervals (16.8%), treatment adherence (11.2%) and nAMD therapy (12.2%). Relatives were particularly interested in legal and social issues and assistive devices (14.3% and 13.2%, respectively).

**Conclusions:** The results indicate that there is a need for an individual care, e.g. provided by a PSP, for patients with retinal diseases. PSPs should be considered a complementary support to the medical treatment or care provided by physicians.

---

**Retinal diseases that lead to visual impairment and loss of vision not only display a medical challenge – they also represent a psychological and emotional burden for patients and their relatives. Patient support programmes (PSPs) can provide tailored, patient-centred support in managing the disease and maintaining treatment adherence. In September 2023, the German telephone-based PSP ‘augenblicke – Das Telefon mit Herz’ was launched for patients with retinal diseases and their relatives**.

Retinal diseases such as neovascular age-related macular degeneration (nAMD), diabetic macular edema (DME) or macular edema due to retinal vein occlusion (RVO) represent a major challenge for patients due to the regular intravitreal injection therapy (IVT) and associated frequent control examinations that are essential for maintaining visual acuity [17, 18]. Both time requirements and unrealistic therapy expectations can negatively impact patient motivation. In everyday care, patients often don’t adhere to or discontinue IVT [23]. One contributing factor is a lack of patient knowledge – however, a comprehensive understanding of the disease is essential for achieving treatment goals [11]. Furthermore, the concurrent management of conditions such as geriatric diseases or diabetic complications, as well as other chronic comorbidities, can pose a challenge [7]. With prolonged or progressive disease, patients’ need for support increases [21]. Accordingly, it is essential to consider social and legal issues such as eligibility for social benefits, transportation costs or exemptions from co-payments.

A diagnosis of a retinal disease requires not only patients to adapt to the new situation. Family members are often heavily involved in organising daily routines and therapy appointments, as well as in providing transportation to these visits. They frequently participate in consultations with the treating physicians, e.g. during discussions of treatment plans [21]. They play a central role by providing emotional support and encouraging adherence to therapy. However, this comprehensive level of involvement can also place a psychological burden on caregivers, who – particularly as the disease progresses – often experience emotions similar to those of patients, e.g. sadness, anxiety, frustration and depression [21].

## PSP – Patient-centred support

The challenges faced by individuals with retinal diseases and their relatives vary from case to case. Disease progression is variable, resulting in unique disease-associated impairments and needs of each patient. In addition to the essential disease-specific patient tailored treatment by the attending physician, patients and their relatives may benefit from holistic, individual and personal support offered by patient support programmes (PSPs).

PSPs are playing an increasingly important role in supporting patients with chronic diseases and their relatives [6]. They represent extended self-management support programmes that aim to offer tailored solutions beyond medication and can improve treatment adherence. In addition to addressing patient-specific questions about the disease and the treatment (necessity) – issues often insufficiently covered in everyday practice – patients can also receive individualised, psychological and emotional support [6, 14]. The telephone-based PSP presented here was specifically developed to support patients with nAMD, DME, and RVO, as well as their relatives. The aim of the PSP is to support patients according to their individual needs, to assist in autonomous disease management and to promote intrinsically motivated adherence to therapy. The collection of health- and therapy-related data within the framework of the PSP is intended to enhance the understanding of patient needs and to contribute to improvements in health care provision and everyday treatment.

## Design and methods

In September 2023, the telephone-based PSP ‘augenblicke – Das Telefon mit Herz’ was initiated for patients with nAMD, DME, and RVO, as well as for their relatives. Patients or their relatives can participate free of charge and independently of the prescribed therapy. They can either contact the programme directly by phone or request a callback via the website https://www.meineaugenblicke.de/. Registration is then completed by phone or, alternatively, participants can register by email using a registration form. After registration, each PSP participant is assigned a personal contact person with professional training in a healthcare profession, who provides regular support calls. During the initial contact, the individual support needs of the patient or relative are determined. The frequency and content of the support calls are tailored to the patients’ needs and life circumstances. The individual calls are not time-limited. Following the telephone calls, participants receive written informational materials covering the topics discussed. Participants also have the option of contacting a free service number in case of any questions.

### Objectives/core elements of the patient support programme

To ensure holistic, patient-centred support, specific content and objectives of the PSP have been defined. These core elements include educational elements (e.g. disease information and the necessity of therapy), as well as psychological and emotional support for the patients. The PSP also offers advice on applying for benefits and assistance with organising treatment appointments (**Figure 1**).

**Figure 1.**
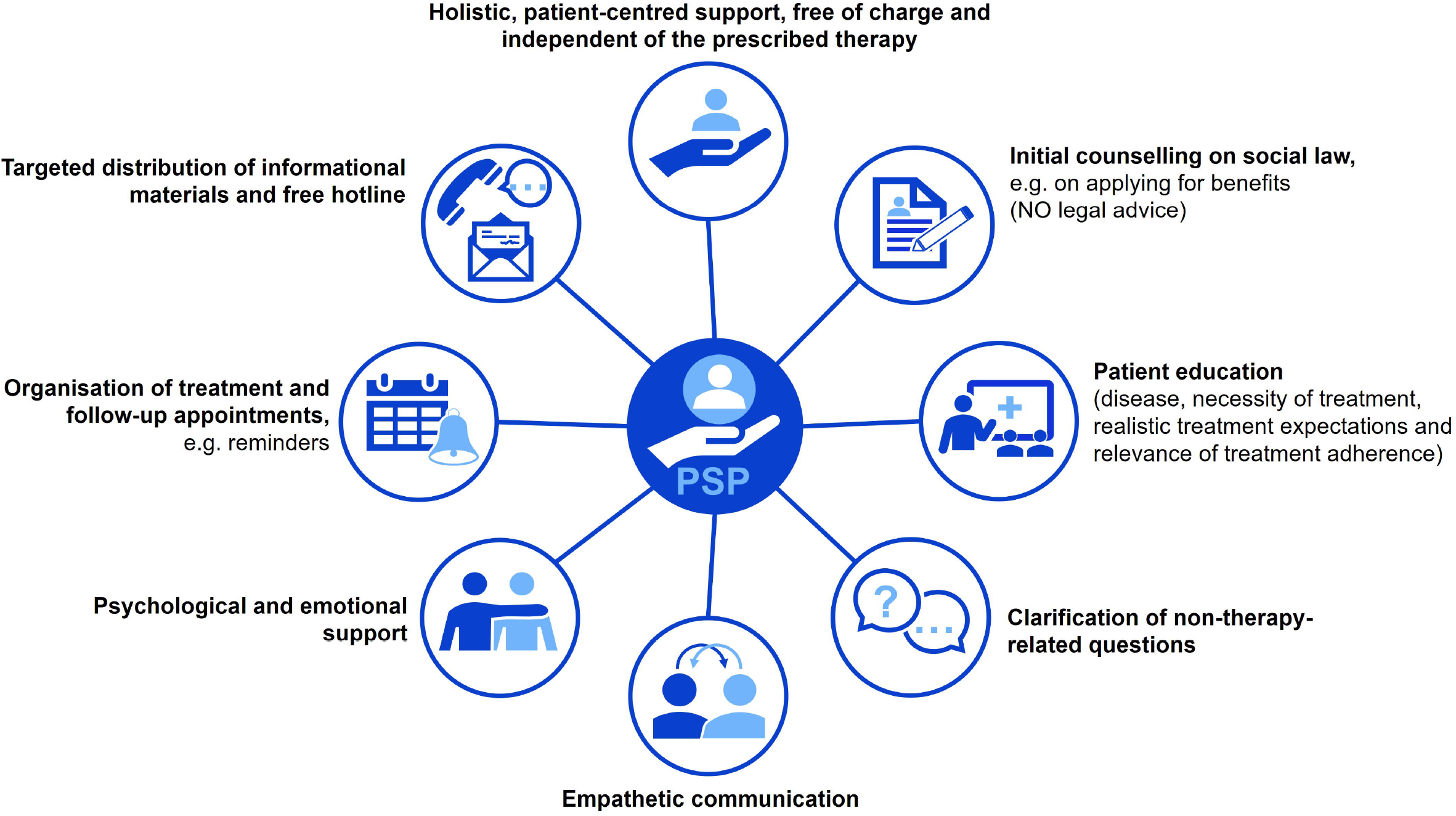
Core elements of the PSP.

### Data collection

By accepting the terms of use, all participants consented to anonymous data analysis. Data collection is performed reactively, with data from the conversations being added to the database successively (no video or audio recordings). No retrospective collection of additional data is conducted. The data are evaluated in anonymised and aggregated form and stored in a password-protected database/data centre (cloud, VPN) operated by dialog4health GmbH in Germany, in compliance with the General Data Protection Regulation (GDPR). In case of an adverse event related to a product of Roche Pharma AG, it is immediately reported to the responsible pharmacovigilance department in compliance with all safety regulations.

## Results

### PSP participants

Since September 2023, a total of 207 participants (188 patients, 19 relatives) have registered for the PSP. At the time of data cutoff (August 31, 2025), 182 patients and 17 relatives continued to participate in the PSP (dropout rate: 3.9%). According to participants’ reports, the majority of all initially registered PSP participants were patients with nAMD (77.8%; N=161), followed by patients with RVO (7.2%; N=15) and DME (6.3%; N=13). Participating relatives were exclusively relatives of nAMD patients (9.2%; N=19) (**Figure 2**).

**Figure 2.**
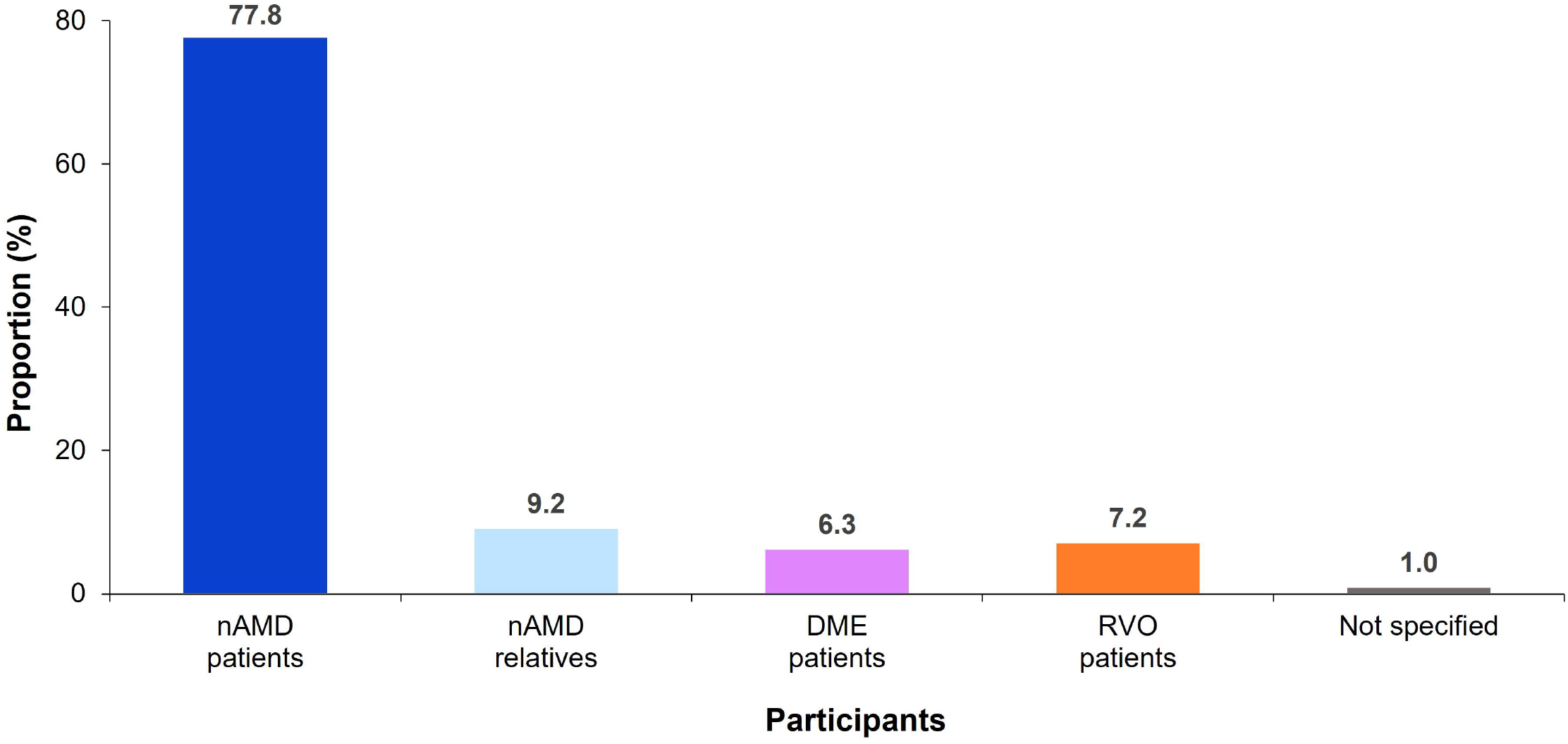
Proportion of participants by indication. The proportion refers to the total number of all initially registered PSP participants (N=207). Three participants each stated two indications (multiple answers possible).

Of all PSP-participating patients (N=188 initially registered), 63.8% provided information about their medication. The most commonly administered drugs included bevacizumab^1^, aflibercept, faricimab and ranibizumab (20.2%, 18.6%, 14.9% and 8.0%, respectively). Twenty-five patients (13.3%) reported not to receive treatment. No information on current therapy was available for approximately 1/4 of the patients (**Figure 3**).

**Figure 3.**
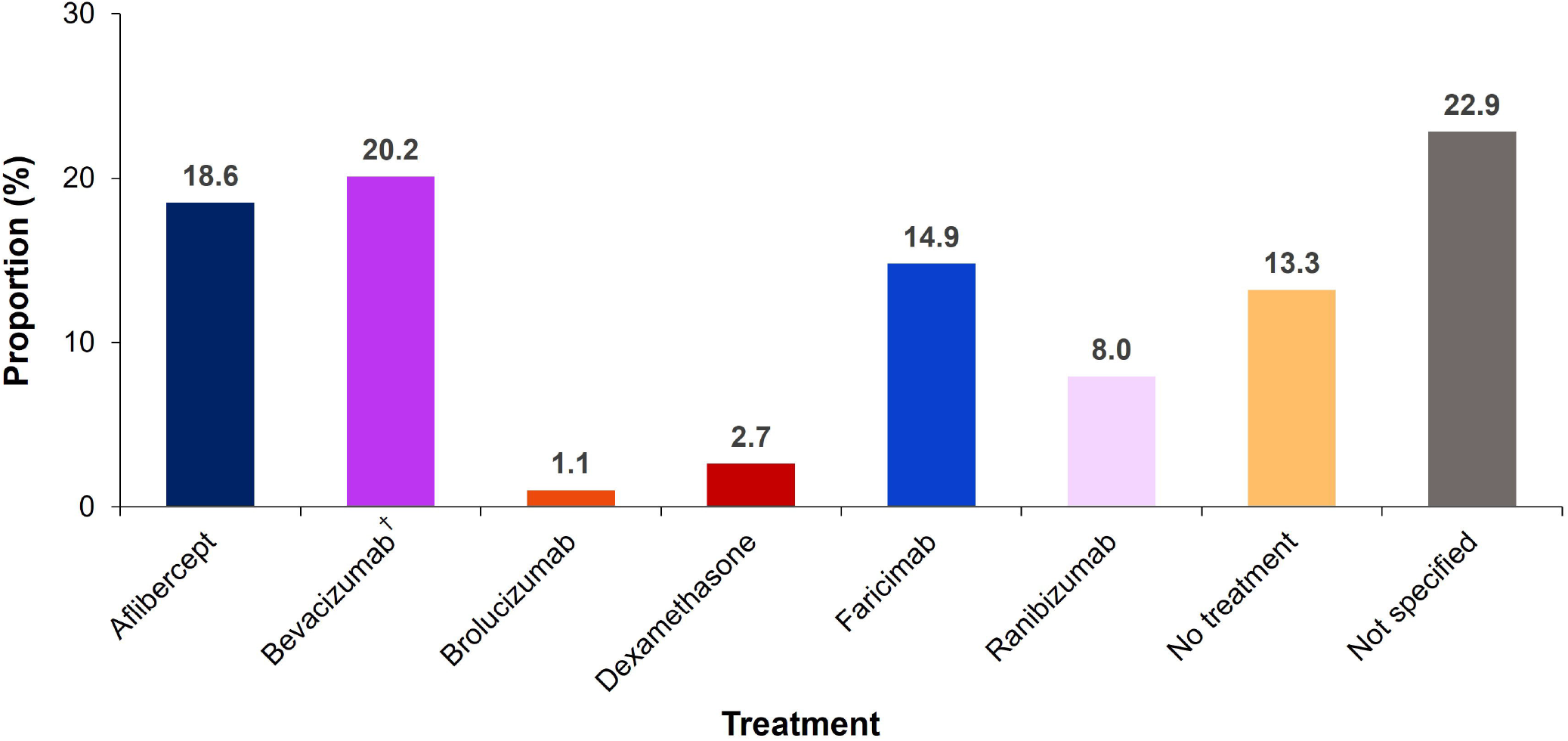
Proportion of patients by therapy. The proportion refers to the total number of all initially registered PSP patients (N=188). Three patients received different intravitreal therapies per eye (multiple answers possible). ^†^Off-label use for the treatment of nAMD, DME, and RVO

### Registration and awareness

The majority of the 207 participants registered for the PSP by telephone (88.0%; N=182). Registration via the website (6.8%; N=14) or the flyer-attached registration form was less common (5.3%; N=11).

Most participants reported having learned about the PSP through print media (65.7%). Additional sources included the PSP website (11.1%) and referrals from their treating physician or nurse (5.8%). Social media or informational materials in the waiting rooms, were rarely the reason for taking note of the PSP (1.9% and 1.0%, respectively) (**Table 1**).

**Table 1.**
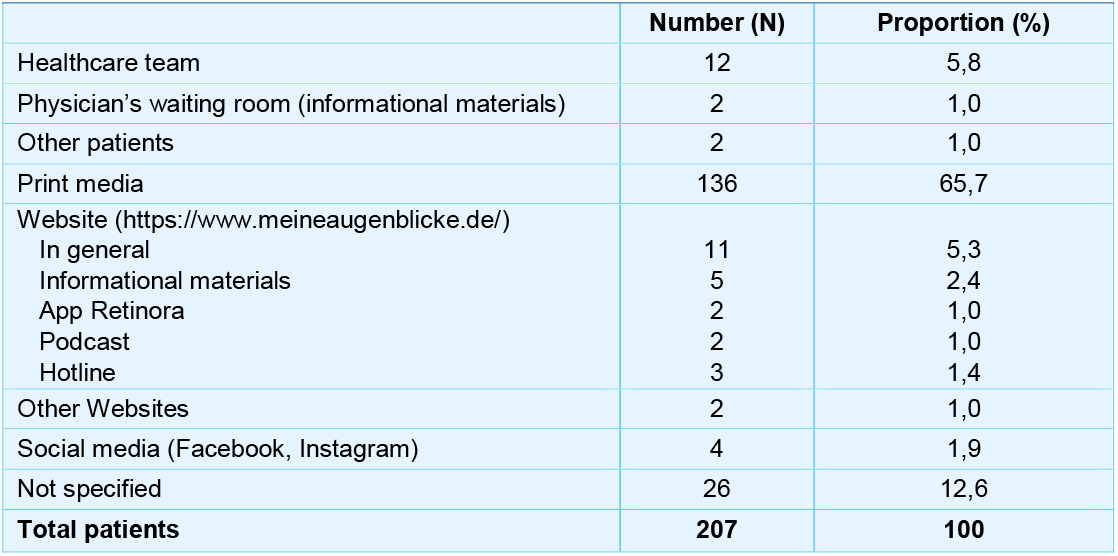
Source of information reported by participants.

| **Table 1. Source of information reported by participants** |  |  |
| --- | --- | --- |
|  | **Number (N)** | **Proportion (%)** |
| Healthcare team | 12 | 5,8 |
| Physician's waiting room (informational materials) | 2 | 1,0 |
| Other patients | 2 | 1,0 |
| Print media | 136 | 65,7 |
| Website (https://www.meineaugenblicke.de/) |  |  |
| In general | 11 | 5,3 |
| Informational materials | 5 | 2,4 |
| App Retinora | 2 | 1,0 |
| Podcast | 2 | 1,0 |
| Hotline | 3 | 1,4 |
| Other Websites | 2 | 1,0 |
| Social media (Facebook, Instagram) | 4 | 1,9 |
| Not specified | 26 | 12,6 |
| **Total patients** | **207** | **100** |

### Contact with participants and key topics

The total number of inbound calls either received via the free service number or personal contact significantly exceeded the number of outbound calls (N=1,121 vs. N=799). The majority of both callers and recipients were patients (89.1%; N=999 and 91.7%; N=733, respectively). The average duration of outbound calls was 19 minutes 48 seconds (maximum: 2 hours 22 minutes 02 seconds). The duration of inbound calls was not documented.

The majority of inbound calls focused on the topic area ‘nAMD diagnosis’ (25.4% and 26.4%, respectively). Key topics within this area included aetiology, pathology and prevalence of the disease, as well as diagnostic procedures and the prognosis for nAMD. Furthermore, patients were most frequently interested in the distinction between dry and wet AMD, as well as in other retinal diseases (i.e., excluding nAMD, DME, and RVO) (19.9% and 19.4% of all patient inbound calls received, respectively). There was also a need for information on treatment-related aspects, such as IVT intervals (16.8%), adherence to IVT (11.2%), and nAMD treatment (12.2%).

In contrast, relatives were more likely to inquire about legal and social issues or assistive devices (14.3% and 13.2% of all inbound calls from relatives, respectively) (**Table 2**).

**Table 2.** Top 5 topics of inbound calls from patients and relatives.

| Top 5 topics* |  | Number (N) | Proportion (%) |
| --- | --- | --- | --- |
| **Inbound calls from patients (N=999)** |  |  |  |
| 1. | nAMD diagnosis | 254 | 25,4 |
| 2. | Dry vs wet AMD | 199 | 19,9 |
| 3. | Other retinal diseases | 194 | 19,4 |
| 4. | IVT intervals | 168 | 16,8 |
| 5. | nAMD treatment | 122 | 12,2 |
| **Inbound calls from relatives (N=91)** |  |  |  |
| 1. | nAMD diagnosis | 24 | 26,4 |
| 2. | nAMD treatment | 13 | 14,3 |
| 2. | Legal and social issues | 13 | 14,3 |
| 4. | Assistive devices | 12 | 13,2 |
| 5. | Dry vs wet AMD | 10 | 11,0 |
| 5. | Other retinal diseases | 10 | 11,0 |

The percentage refers to the total number of inbound calls from patients or relatives (N=999 and N=91, respectively). \*Multiple answers possible. Compliance with the obligation to report suspected adverse reactions was ensured.

## Discussion

The retinal diseases nAMD, DME, and RVO display a common cause of severe vision loss, which, if left untreated, can lead to blindness [9, 20, 22]. Current standard treatment for these conditions involves regular intravitreal anti-VEGF therapy [1, 15, 19]. However, the frequent intravitreal injections required to maintain visual acuity may pose a significant burden for both patients and their relatives [17, 18, 19].

To support affected individuals in this situation, a telephone-based PSP was launched in September 2023. It was developed for patients with nAMD, DME, and RVO and their relatives, with the aim of providing holistic, patient-centred support. Among other things, this support is intended to encourage motivation to continue the time-intensive treatment. Treatment continuation, i.e. strict adherence to the often highly frequent injection appointments prescribed by the physician, is critical for maintaining visual improvement and preventing further vision loss. It has been shown previously that the number of injections received is directly related to the improvement or maintenance of visual acuity and, along with therapy monitoring, is partly responsible for the visual outcome [8]. Consequently, non-adherence or delays in scheduled appointments negatively impacts visual acuity in nAMD patients [13]. Despite the severe consequences of non-adherence, German real world data demonstrated that as early as 3 months after initiating IVT, one-third of the patients with nAMD had not received any treatment or follow-up for at least 6 weeks. After 6 months, this even applied to two-thirds of patients [4].

The reasons for poor adherence of patients are diverse and individual. Factors as advanced age or lower baseline visual acuity are associated with non-adherence [5, 16]. A good response appears to have an even greater impact on adherence than baseline visual acuity [11]. On the other hand, unrealistic therapy expectations have led patients to question treatment effectiveness, potentially leading to a loss of motivation to continue treatment [5, 11]. Moreover, patients often lack sufficient knowledge about their disease and the necessity for regular intravitreal injections and follow-up examinations [11]. This PSP also revealed a clear need for information among participating patients regarding their own retinal disease and therapy (e.g., injection intervals and adherence to IVT, nAMD treatment).

A well-informed patient is a key prerequisite for intrinsically motivated treatment adherence [11]. It is therefore crucial to consider modifiable risk factors, such as patient knowledge, by providing comprehensive information about the disease and the need for therapy as well as communicating realistic therapy expectations. PSPs can make a valuable contribution by addressing various patient-relevant aspects. The assigned contact person can respond to patient questions using patient-friendly, accessible language, build trust, motivate patients, and reduce fears and concerns shared during the conversation. Furthermore, patients can be reminded of their treatment and follow-up appointments, and social law issues can be addressed.

PSPs are already being applied in various indications and have the potential to improve care for chronic diseases requiring complex treatment regimens [6]. Intensive patient care, e.g., within PSPs, has been shown to increase adherence in rheumatoid arthritis, type 2 diabetes mellitus, multiple sclerosis, and chronic myeloid leukemia [2, 10, 12, 24]. A retrospective study of patients with chronic diseases also demonstrated that PSP participation can have a positive effect on adherence. In this study, PSP participation was associated with a 29.3% higher adherence rate and a 22% lower treatment discontinuation rate compared to non-participation [2]. Patients with retinal diseases also benefit from a PSP: In an Australian study, nAMD patients who received intravitreal therapy and participated in a PSP demonstrated significantly higher persistence compared to patients without PSP support (after 12 months: 93% vs. 76%; after 24 months: 88% vs. 64%) [3].

Given the limited time resources of physicians and the increasing financial pressure on practices and treatment centres, there is often insufficient time during consultations to address therapy-related and medical issues in detail and iteratively. Clarifying non-therapeutic issues prior to a physician’s appointment may ease the clinical workflow, as it allows the treating physician to focus on therapy-specific topics during the consultation. It is important to note that, therapeutic recommendations are not permitted within the PSPs. However, PSPs offer the opportunity to address topics that go beyond medical information. Even though PSPs can neither replace nor intent to replace medical care by a physician, they may serve as a valuable complement to medical treatment and physician support, closing potential gaps in patient care and support.

### Limitations

In the PSP presented here, data are not collected systematically (e.g., no use of validated questionnaires or documentation of demographic data). Data collection is reactive and individualised based on the content of the conversation between participants and PSP contact persons. Patients can participate in the PSP at any time, resulting in varying participation durations. Due to the non-systematic nature of data collection, longitudinal follow-up is not uniformly possible. Furthermore, strict efficacy parameters are not recorded. A potential bias cannot be ruled out, as only a specific type of patient may seek support from the PSP. Nevertheless, a valuable insight was gained into the areas in which patients and their relatives individually need support.

#### Practical implications

- Regular use of the PSP and the topics covered in the most frequently asked questions indicate that patients with retinal diseases need individualised support through such programmes.
- PSPs should be regarded as complementary to medical treatment and care provided by physicians.
- Increased awareness among physicians and patients could help expand access to patient programmes like this one and enable comprehensive patient care – including assistance for the healthcare team.

#### Compliance with ethical guidelines

By accepting the terms of use, all participants have consented to anonymous data analysis. This article does not include any studies involving humans or animals.

#### Potential conflicts of interest

Kai Januschowski: Roche (speaker/research/consulting activities); Zeiss, Oertli (consulting activities)

Eva-Maria Konrad: Roche (speaker/consulting activities, travel expenses)

Anna Thonet: No conflict of interest

Florian Karsch: The setup and implementation of the PSP by dialog4health was financed by Roche Pharma AG.

Virginia Ziska: Roche Pharma AG (employee and stock owner)

Kristina Gelblin: Roche Pharma AG (employee and stock owner)

## Data availability

Raw data were generated at Roche Pharma AG, Grenzach-Wyhlen, Germany. The data are not publicly available due to restrictions regarding data protection, as they include information that could compromise the privacy of research participants. Anonymised data supporting the findings of this study are available on request from.

## Acknowledgements

We would like to thank Dr Sandra Blümich and Dr Eva-Maria Wagner, employees of Roche Pharma AG, for their support throughout the project. We would also like to thank the patient organisation PRO RETINA Deutschland e.V. for valuable discussions during the planning and design of the PSP to meet the needs of patients and their relatives, as well as for possible approaches to solutions. We would also like to thank the patient and relative panel, consisting of patients with nAMD/DME/RVO and their relatives, for their input into the design of this PSP. Medical writing assistance of the manuscript was carried out by med:unit GmbH, Rheinbach, and was funded by Roche Pharma AG. The authors had full editorial control at any timepoint and gave their final approval.

## Footnotes

1 Off-label use for the treatment of nAMD, DME, and RVO

## Notes

### Author Declarations

We confirm that data collected as part of the PSP consist of anonymised individual-level data from participants, which have been evaluated in aggregate form.

